# T Cell Positive B Cell Negative Flow Cytometry Crossmatch (FCXM): Frequency, HLA-Locus Specificity, and Mechanisms Among 3073 Clinical FCXM Tests

**DOI:** 10.1101/2021.05.20.21257541

**Authors:** Prabhakar Putheti, Vijay K Sharma, Rex Friedlander, Arvind Menon, Darshana Dadhania, Thangamani Muthukumar, Manikkam Suthanthiran

## Abstract

**Background:** A T cell positive and B cell negative (T+B-) flow cytometry crossmatch (FCXM) result remains a conundrum since HLA-class I antigens are expressed on both T and B cells. We investigated the frequency, HLA specificity of the antibodies and mechanisms for the T+B- FCXM result.

**Methods:** We analyzed 3073 clinical FCXM tests performed in an American Society of Histocompatibility and Immunogenetics accredited histocompatibility laboratory. The sera associated with the T+B- FCXM were also tested for donor HLA IgG antibodies using LABScreen™ single antigen assays.

**Results:** Among the 3073 FCXM tests, 1963 were T-B-, 811 were T-B+, 274 were T+B+, and 25 were T+B-. IgG antibodies directed at donor HLA-A, B, or Cw locus determined antigens (DSA) were identified in all 25 sera and the summed mean fluorescence intensity (MFI) of DSA ranged from 212 to 53,187. Correlational analyses identified a significant association between the summed MFI of class I DSA, and the median channel fluorescence (MCF) of T cells treated with the recipient serum (Spearman rank correlation, r_s_=0.34, P=0.05) but not with the MCF of B cells (r_s_=0.23, P=0.24). We identified that differential binding of anti-HLA antibodies to T cells and B cells and the B cell channel shift threshold used to classify a B cell FCXM are potential contributors to a T+B- FCXM result.

**Conclusions:** Our analysis of 3073 FCXM, in addition to demonstrating that HLA antibodies directed at HLA-A, B or Cw locus are associated with a T+B- result, identified mechanisms for the surprising T+B- FCXM result.

## INTRODUCTION

Kidney transplantation is the treatment of choice for most patients with irreversible kidney failure, but pre-sensitization to donor human leukocyte antigens (HLA) and antibody-mediated rejection, undermine its full benefits. In a landmark publication, Patel and Terasaki reported that 24 of 30 kidneys transplants performed across a positive complement dependent cytotoxicity crossmatch (CDC-XM) failed in an accelerated fashion^1^. Test systems to detect circulating antibodies at a higher sensitivity than CDC-XM have been developed, and the flow cytometry crossmatch (FCXM) is in widespread use to identify donor specific IgG antibodies (DSA)^2-5^. Pre-transplant FCXM results are used in many centers for optimizing donor selection, and pre-transplant T and B cell FCXM results have been associated with allograft outcomes^6-10^. FCXM detects binding of any type of antibodies to T cells and B cells including autoantibodies and non-HLA antibodies.

HLA-class I antigens are expressed on T cells and B cells, and some but not all studies suggest that the cell surface expression of HLA-class I antigens is higher on B than T cells^11,12^. Thus, a T+B+ FCXM, rather than a T+B-, is the expected result in the presence of HLA-class I antibodies. A T-B+ FCXM is the anticipated result in the presence of HLA-class II antibodies only since HLA-class II antigens are expressed on B cells but not on resting T cells^13^.

We aimed to determine the frequency, HLA-specificity of the antibodies and potential mechanisms responsible for the T+B- FCXM. We analyzed 3073 clinical FCXM tests performed using a standardized protocol in an American Society of Histocompatibility and Immunogenetics accredited Histocompatibility Laboratory and report here the frequency, HLA-specificity of the antibodies and mechanisms responsible for the T+B- FCXM.

## MATERIALS AND METHODS

### Isolation of mononuclear cells for FCXM test

Peripheral blood collected in acid citrate dextrose tubes was used for isolation of peripheral blood mononuclear cells (PBMC) using Ficoll-Paque™ Premium solution (GE Healthcare Biosciences AB, Pittsburgh, PA) density gradient centrifugation. Mononuclear cells from the lymph node and spleens from deceased donors were also isolated using Ficoll-Paque™ Premium solution density gradient centrifugation. The mononuclear cells from the interface were washed thrice with phosphate buffered saline (PBS) and the red blood cells were lysed with tris ammonium chloride. PBMC were counted using either Neubauer counting chamber or LUNA™ Automated Cell Counter (Model# L20001; Logos Biosystems, Inc., Annandale, VA). Cell viability was assessed using Trypan blue exclusion test or acridine orange/propidium iodide assay. The viability of mononuclear cells exceeded 80% in the assays reported here.

### Separation of serum

Peripheral blood collected in serum separation tubes was centrifuged at 2300 revolutions per minute for 20 minutes. The serum was collected and treated with 0.3% sodium azide prior to storage.

### FCXM

FCXM was performed in a 96 well V-bottom plate (Cat#3894, Corning, Inc., Corning, NY). 300,000 PBMC or mononuclear cells, isolated from the spleen or the lymph node, were incubated with 30 µl of negative control serum (NCS) (normal male human AB serum free of HLA-class I and II antibodies); recipient serum (RS); or positive control serum (PCS, pooled serum from highly sensitized patients with broad HLA reactivity) for 20 minutes at 22°C. Prior to testing, the NCS, the RS and the PCS were centrifuged at 42,030 relative centrifugal force to remove any aggregates or cryoprecipitate.

The mononuclear cells used as target cells in the FCXM were washed twice with wash buffer (PBS with 0.1% FBS and 0.1% Sodium Azide) and stained with 10 µl each of anti-CD3 PerCP mAb (Cat#340663; BD Biosciences, San Jose, CA); anti-CD19 PE mAb (Cat#340720; BD Biosciences); and fluorescein (FITC) affinity purified F (ab’) 2 fragment goat anti-human IgG, Fc□fragment specific (Cat# 109-096-098; Jackson ImmunoResearch Laboratories, Inc.; 1:200 diluted) for 30 minutes at 4°C. The cells were then washed once, fixed with 1% paraformaldehyde, and FCXM was performed using BD FACS Canto II instrument and the data were analyzed using the FACSDiva software (versions 7.1.3 and 8.0.1). Lymphocytes were gated using forward scatter (FSC) versus side scatter (SSC) plot. Median channel fluorescence (MCF) of goat-anti-human IgG FITC fluorescence on T cells and on B cells was measured from the histograms. Channel shift (CS) was calculated by subtracting the MCF observed with NCS from the MCF observed with the RS or PCS.

A T cell FCXM test was reported as positive if the MCF of T cells incubated with the RS showed a 40 CS or higher. A B cell FCXM test was reported as a positive test if the MCF of B cells incubated with the RS showed a 50 CS or higher. The CS thresholds were based on the interquartile range (IQR, calculated by subtracting the 25^th^ percentile value from the 75^th^ percentile value) observed in our laboratory following treatment of target mononuclear cells with NCS. In 2020 consecutive T cell FCXM performed at our center, the median MCF of T cells treated with NCS was 229 and the IQR was 39, and in 2019 consecutive B cell FCXM, the median MCF of B cells treated with NCS was 252 and the IQR was 47. In eight consecutive American Society for Histocompatibility and Immunogenetics (ASHI)-proficiency challenges involving 66 unique sera and 16 unique HLA-typed cells, the use of these CSs, 65 of 66 T cell FCXM tests and 64 of 66 B cell FCXM were concordant with the 80% ASHI consensus result (ground truth).

### Detection of Anti-HLA Antibodies

Sera, untreated or treated with EDTA (0.025M final concentration), were tested for the presence of anti-HLA IgG antibodies leveraging Luminex^®^ bead-based multiplexing technology. Anti-HLA-class I HLA-A, B, C IgG antibodies and anti-HLA-class II HLA-DR, DQ, DP IgG antibodies were identified using LABScreen™ Single Antigen class I assay (LS1A04) and LABScreen™ Single Antigen class II assay (LS2A01), respectively (One Lambda, Inc., Canoga Park, CA). In the LABScreen Single Antigen (LSA) assay, 10 µl of serum samples were incubated with 2.5 µl of microbeads coated with HLA-class I or HLA-class II antigens, respectively, at 22°C for 30 minutes. The specimens were then washed thrice in 1X wash buffer (One Lambda, Inc.). The microbeads were treated with PE labeled goat-anti-human IgG polyclonal antibody (Cat#LS-AB2, One Lambda, Inc.) at 22°C for 30 minutes in dark and washed thrice in wash buffer. The LABScan 100/200 (Luminex Corporation, Austin, TX) was used for acquisition of microbead fluorescence and data analysis. Data were then exported to HLA fusion software (One Lambda, Inc.) for analysis.

### Data Analysis

A total of 3609 FCXM tests were performed in our laboratory from February 2014 to April 2018. We report data from 3073 FCXM tests using potential organ donor’s cells as target cells, and after exclusion of recipient auto FCXM and repeat donor FCXM with the same result.

In the Lumine single antigen bead (LSA) assay, any single antigen bead with a trimmed mean fluorescence intensity (MFI) of ≥2000 was scored as positive for the presence of anti-HLA IgG DSA in accord with our threshold used in the ASHI Proficiency challenges. We summed MFI of class I and MFI class I and II for the correlational analyses between summed MFI and MCF of T cells and B cells treated with RS, respectively.

### Statistical Analysis

Statistical analysis was performed using Prism 9 (GraphPad Software, San Diego, CA, USA). We summarized the data as median and IQR and used Mann-Whitney test and Wilcoxon matched-pairs signed-rank test to compare the groups. A two-sided P-value <0.05 was considered significant. Data analysis was performed under Weill Cornell Medicine institutional review board approved protocol.

## RESULTS

### Frequency of T+B- FCXM Result

A total of 3609 clinical FCXM tests were performed in our laboratory from February 2014 to April 2018 and data from 3073 FCXM tests were included in downstream data analysis (Figure 1). Among the 3073 tests, 1963 FCXM tests were negative (64%) and 1110 FCXM tests were positive (36%). Among the positive tests, 811 were T-B+ (73%), 274 were T+B+ (24.7%) and 25 FCXM were T+B- (2.3%).

**Figure 1.**
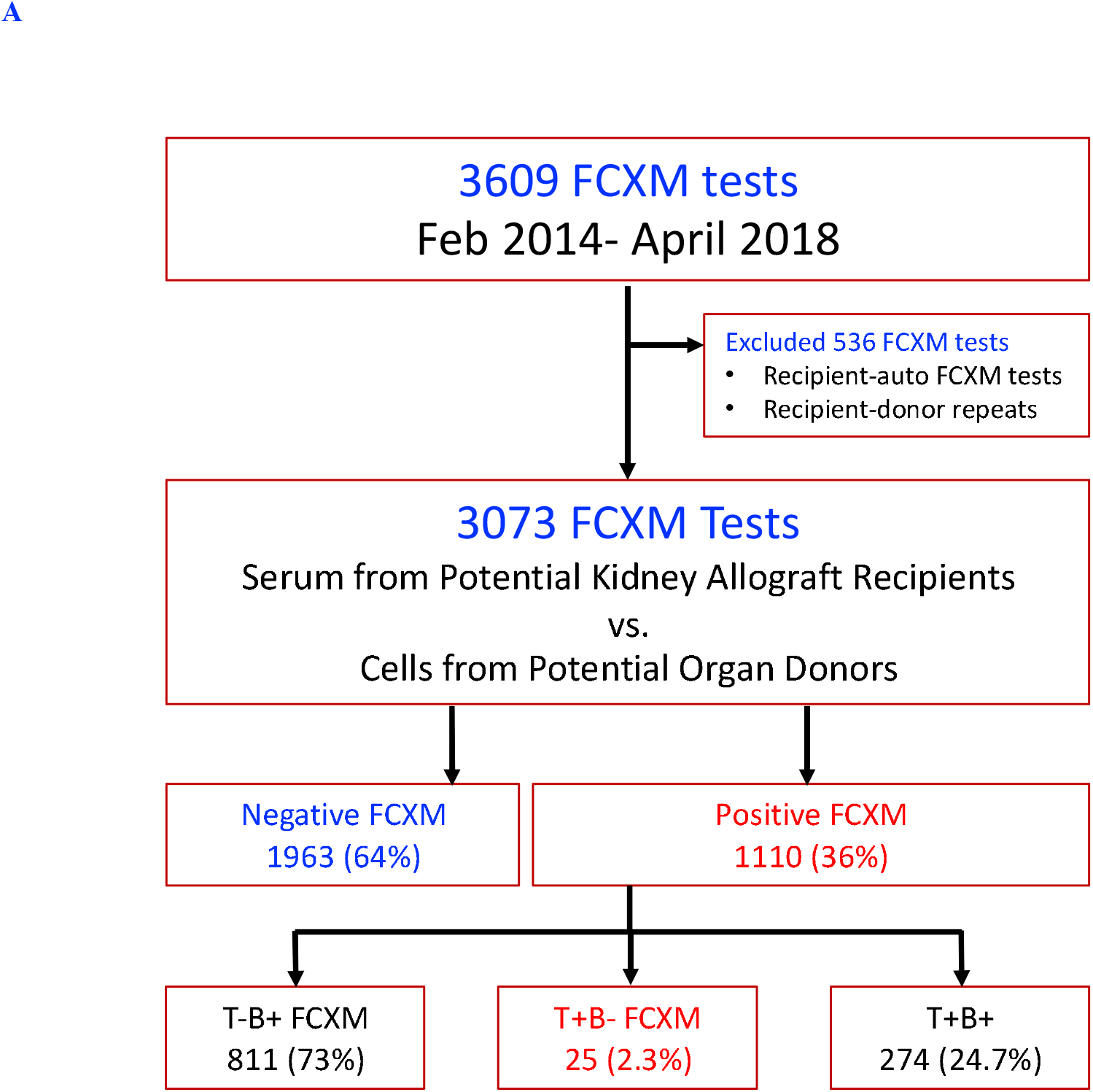
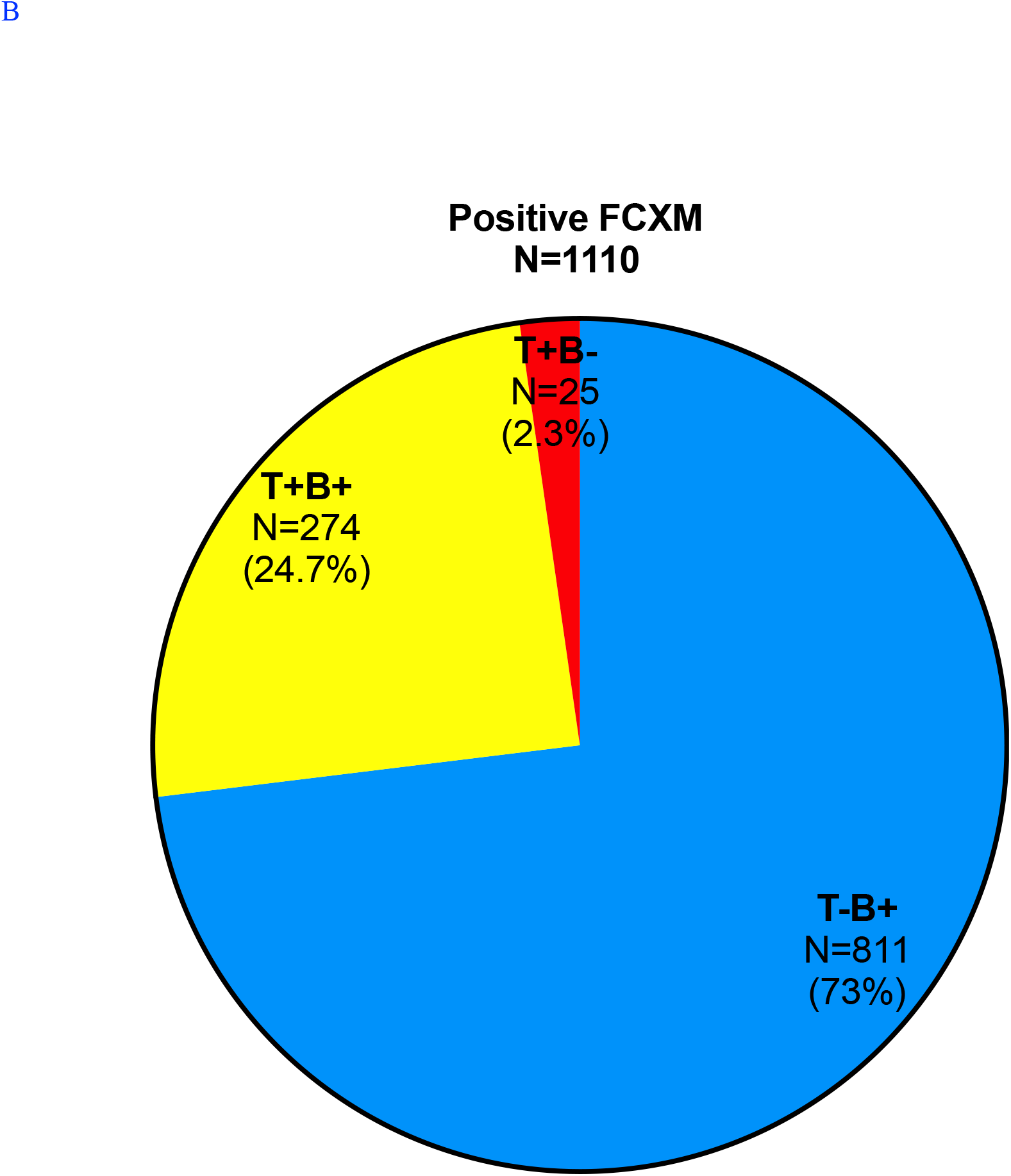
Frequency and distribution of flow cytometry crossmatch test results. (A) A total 3609 clinical flow cytometry crossmatch (FCXM) tests were performed from February 2014 to April 2018 at the Immunogenetics and Transplantation Center (IGT), an ASHI-accredited histocompatibility laboratory. The crossmatches were performed for selecting a compatible kidney donor. Auto FCXM test and repeat donor FCXM tests using the same serum/ donor cell combination were excluded and 3073 donor FCXM tests were included for downstream data analysis. Among these 3073 FCXM tests, 1963 were reported as negative FCXM (63.9%) and the remaining 1110 were reported as FCXM positive (37.1%) tests. (B) Among the positive 1110 FCXM tests, 811 were T–B+ (73%), 274 were T+B+ (24.7%) and 25 were T+B- (2.3%).

24 unique patients awaiting kidney transplantation contributed the 25 sera associated with the T+B- and one patient’s serum was tested against two different donors and the results were T+B- in both instances. In 12 FCXM, a recent serum and a historical serum from the same patient were tested against the same donor and data analysis showed that 10 of the 12 historical sera were also T+B- (with 2 sera being borderline T positive with CS of 31 and 39) and two were T+B+.

Eleven of the 25 T+B- FCXM (44%) were performed using deceased donor cells as targets and 14 (56%) were performed using living donor cells as targets. PBMC were the targets in 22 of 25 T+B- FCXM, and mononuclear cells isolated from spleen (n=2) or lymph node (*n*=1) were the targets in 3.

### Donor HLA Antibodies Associated with the T+B- FCXM

Donor HLA specific IgG antibodies directed at HLA class I antigens (>2000 MFI) were detected in 23 of 25 sera associated with the T+B- FCXM. In # Case 24, the MFI of IgG DSA directed at donor HLA-A1 was 1403 and the MFI of DSA directed at HLA-B57 was 1741. In Case #25, the highest MFI of DSA was 212. Analysis for HLA-class I locus specificity showed that the antibodies were often directed at more than one locus (Table 1). Among the 23 sera, antibodies directed at HLA-A were identified in12, antibodies at HLA-B in 10, and antibodies at HLA-Cw in 10. Antibodies directed at HLA-class II antigens were also detected in 6 sera.

**Table 1.**
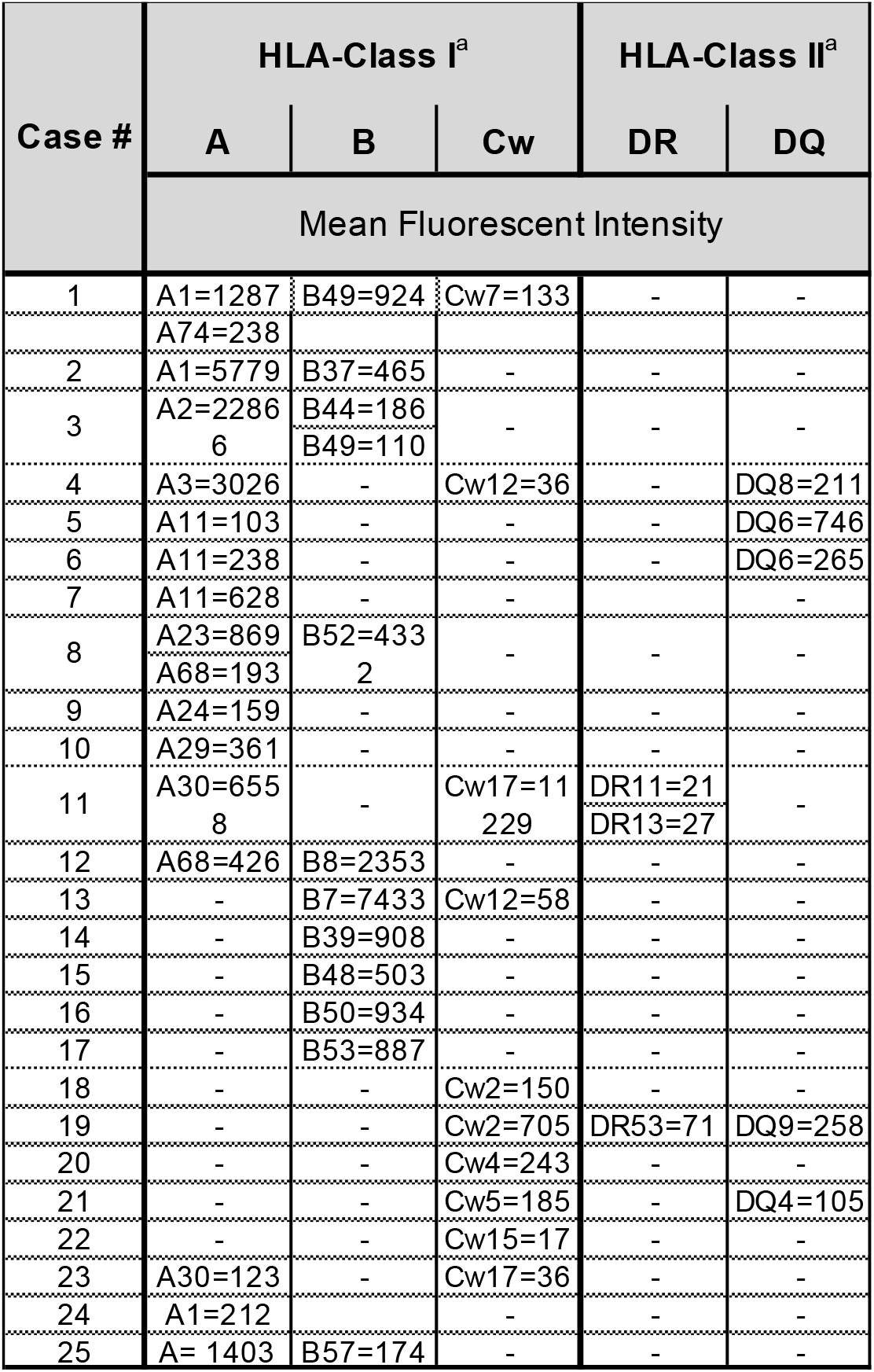
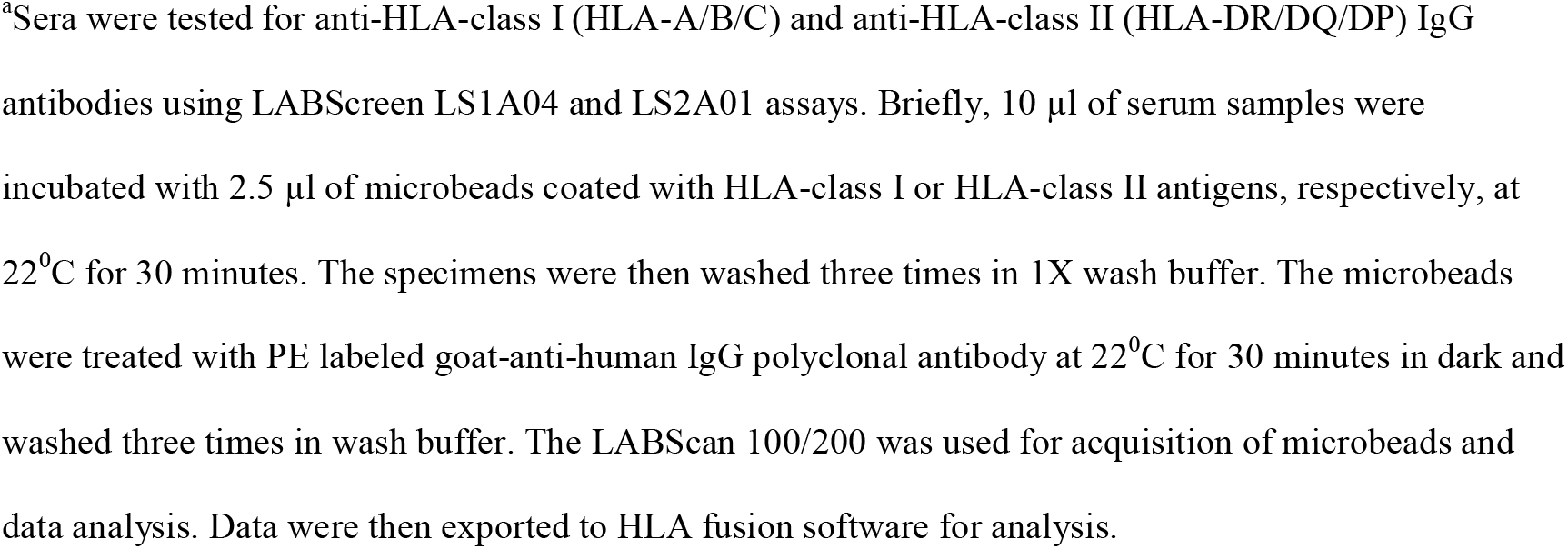
Donor HLA antibodies Associated with T+B- FCXM test result.

| Case # | HLA-Class I<sup>a</sup> |  |  | HLA-Class II<sup>a</sup> |  |
| --- | --- | --- | --- | --- | --- |
|  | A | B | Cw | DR | DQ |
|  | Mean Fluorescent Intensity |  |  |  |  |
| 1 | A1=1287<br>A74=238 | B49=924 | Cw7=133 | - | - |
| 2 | A1=5779 | B37=465 | - | - | - |
| 3 | A2=2286<br>6 | B44=186<br>B49=110 | - | - | - |
| 4 | A3=3026 | - | Cw12=36 | - | DQ8=211 |
| 5 | A11=103 | - | - | - | DQ6=746 |
| 6 | A11=238 | - | - | - | DQ6=265 |
| 7 | A11=628 | - | - | - | - |
| 8 | A23=869<br>A68=193 | B52=433<br>2 | - | - | - |
| 9 | A24=159 | - | - | - | - |
| 10 | A29=361 | - | - | - | - |
| 11 | A30=655<br>8 | - | Cw17=11<br>229 | DR11=21<br>DR13=27 | - |
| 12 | A68=426 | B8=2353 | - | - | - |
| 13 | - | B7=7433 | Cw12=58 | - | - |
| 14 | - | B39=908 | - | - | - |
| 15 | - | B48=503 | - | - | - |
| 16 | - | B50=934 | - | - | - |
| 17 | - | B53=887 | - | - | - |
| 18 | - | - | Cw2=150 | - | - |
| 19 | - | - | Cw2=705 | DR53=71 | DQ9=258 |
| 20 | - | - | Cw4=243 | - | - |
| 21 | - | - | Cw5=185 | - | DQ4=105 |
| 22 | - | - | Cw15=17 | - | - |
| 23 | A30=123 | - | Cw17=36 | - | - |
| 24 | A1=212 | - | - | - | - |
| 25 | A= 1403 | B57=174 | - | - | - |

<sup>a</sup>Sera were tested for anti-HLA-class I (HLA-A/B/C) and anti-HLA-class II (HLA-DR/DQ/DP) IgG antibodies using LABScreen LS1A04 and LS2A01 assays. Briefly, 10 µl of serum samples were incubated with 2.5 µl of microbeads coated with HLA-class I or HLA-class II antigens, respectively, at 22<sup>0</sup>C for 30 minutes. The specimens were then washed three times in 1X wash buffer. The microbeads were treated with PE labeled goat-anti-human IgG polyclonal antibody at 22<sup>0</sup>C for 30 minutes in dark and washed three times in wash buffer. The LABScan 100/200 was used for acquisition of microbeads and data analysis. Data were then exported to HLA fusion software for analysis.

### Characteristics of T+B- FCXM

Table 2 lists the characteristics of the 25 T+B- FCXM. The median (IQR) of MCF was 230 (36) with the potential donor T cells treated with NCS and 250 (65) for the corresponding B cells (*P*<0.05, Wilcoxon signed-rank test for paired samples). This difference remained significant even after exclusion of Case #5 with a low T cell MCF and an unusually high B cell MCF. B cell MCF being higher than T cell MCF was also observed in the 3048 FCXM tests that were not T+B- (B cell MCF=253[45] vs. T cell MCF=233[38], *P<*0.0001).

**Table 2.**
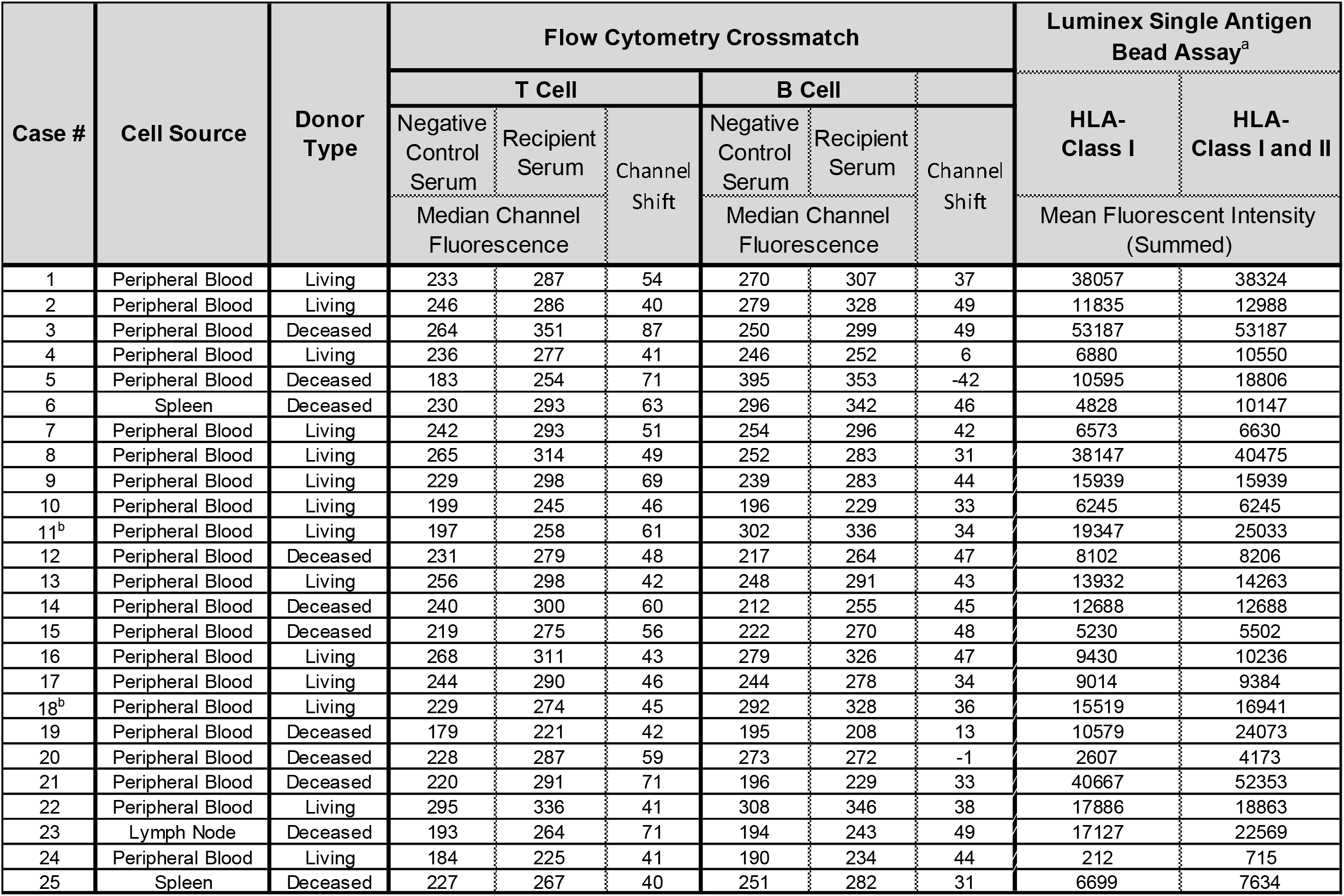
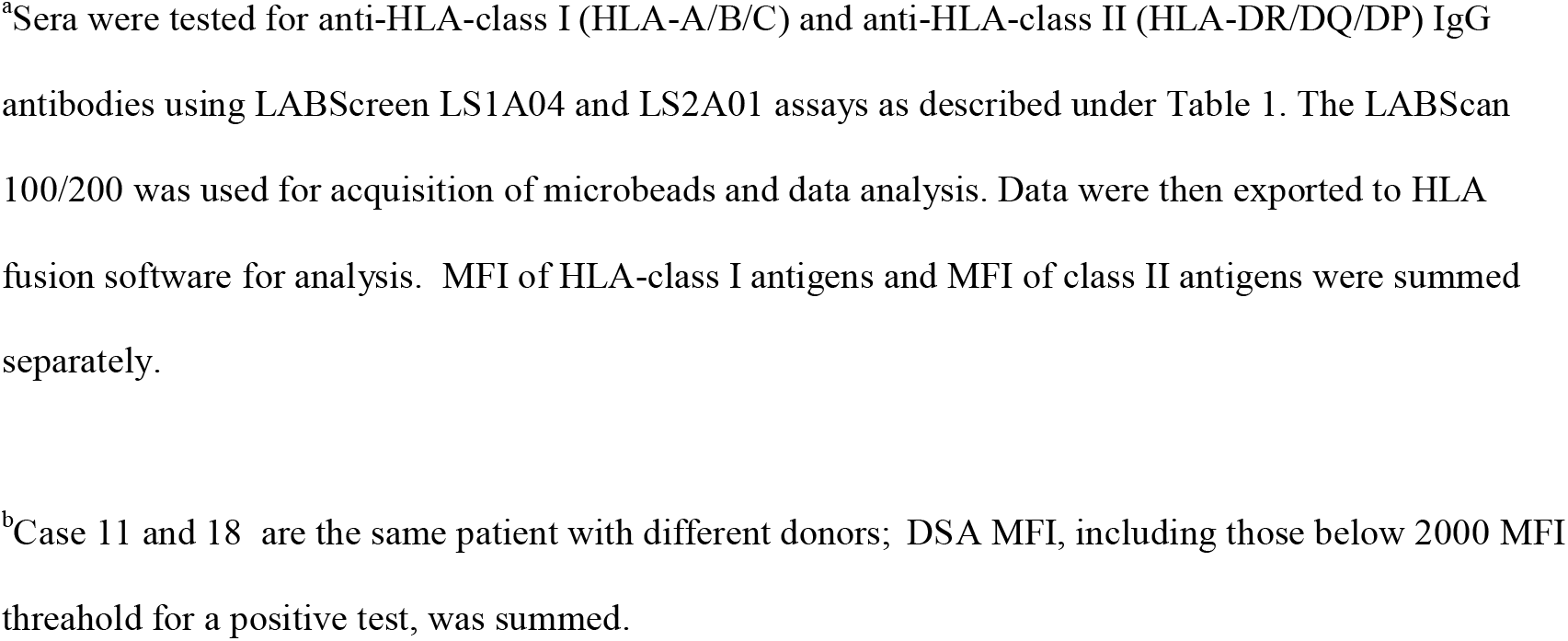
Charecterstics of T+B– FCXM tests.

MCF of T cells and the MCF of B cells treated with the RS in the 25 T+B- FCXM were not different. The median (IQR) MCF was 287(33) with the T cells treated with the RS and 283 (74) for the corresponding B cells treated with the RS (P=0.81).

The channel shift, calculated by subtracting the MCF of cells treated with NCS from the MCF of cells treated with the RS, differed significantly between the T cells and the B cells in the 25 T+B- FCXM. The median (IQR) CS with T cells was 49 (20) and higher than the median (IQR) 38 (15) CS with B cells (P<0.0001).

Figure 2 shows the differential relationship between summed MFI of DSA and the MCF of T cells and MCF of B cells treated with the RS in the 25 T+B- FCXM. A significant and positive association was observed between the summed MFI of class I DSA and T cell MCF treated with RS (Figure 2A, Spearman rank correlation coefficient r_s_=0.34, P=0.05) and there was no significant association between B cell MCF and the summed MFI of class I DSA (Figure 2B, r_s_=0.24, P=0.22) or the summed MFI of class I and class II DSA (Figure 2C, r_s_=0.23, P=0.24).

**Figure 2.**
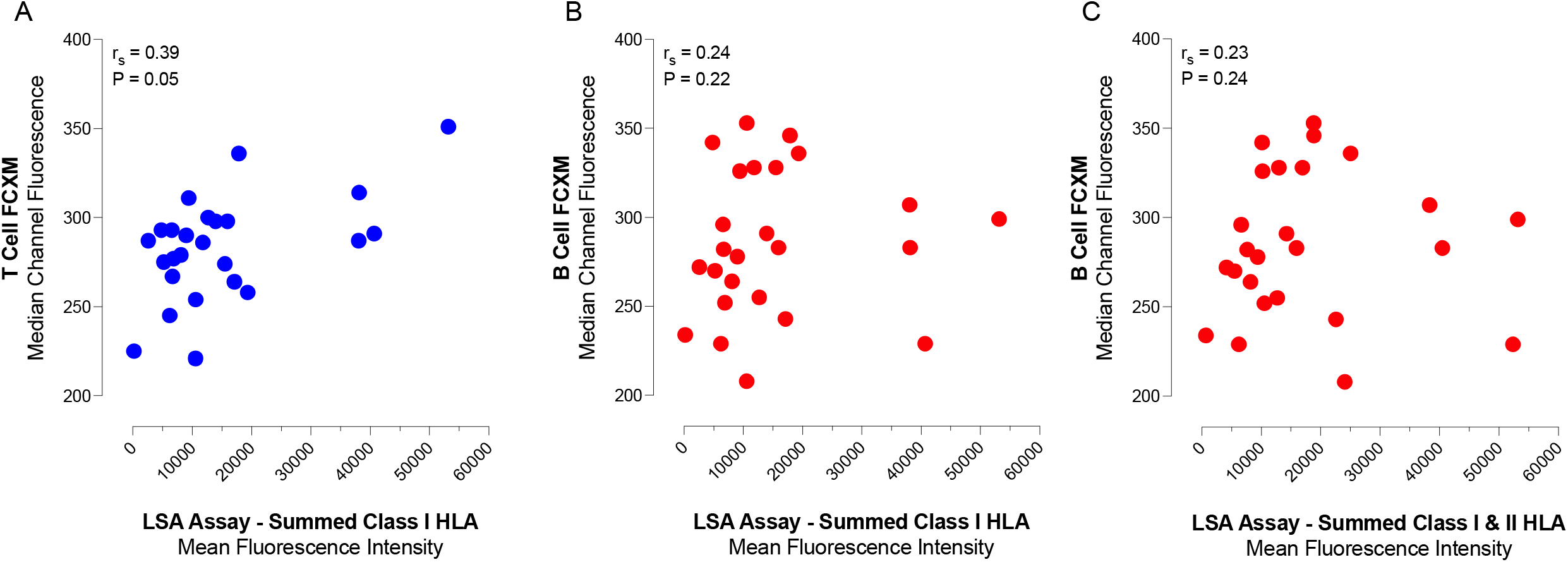
Correlational analysis of summed MFI of donor HLA antibodies in recipient’s serum and the MCF and T cells and B cells treated with the recipient serum. Donor HLA antibodies were identified using the LSA assay and the MFI was summed. (A) A significant and positive association was observed between the summed MFI of class I DSA and the MCF of T cells treated with RS; (B) There was no significant association between the MCF of B cells treated with RS and the summed MFI of class I DSA; and (C) There was no significant association between the MCF of B cells treated with RS and the summed MFI of class I and class II DSA. Spearman rank correlation coefficient r_s_ and associated P values were calculated are included in each panel.

### Potential Mechanisms for the T+B- FCXM Result

A mechanism for the T+B- FCXM is that the MCF of B cells treated with the NCS was uniquely higher when the result is T+B- FCXM compared to the B cell MCF when the result is T+B+ FCXM or T-B+ FCXM. To investigate this possibility, we compared the MCF of B cells treated with NCS among the FCXM classified as T+B-, T+B+, or T-B+ FCXM. The median (IQR) of B cell MCF was 250 (65) in the 25 T+B- FCXM tests, 248 (49) in the 274 T+B+ FCXM, and 254 (45) in the 811 T-B+ FCXM (P=0.2, Kruskal-Wallis test) (Figure 3A). Thus, a uniquely high B cell MCF was not responsible for the observed T+B- test result.

**Figure 3.**
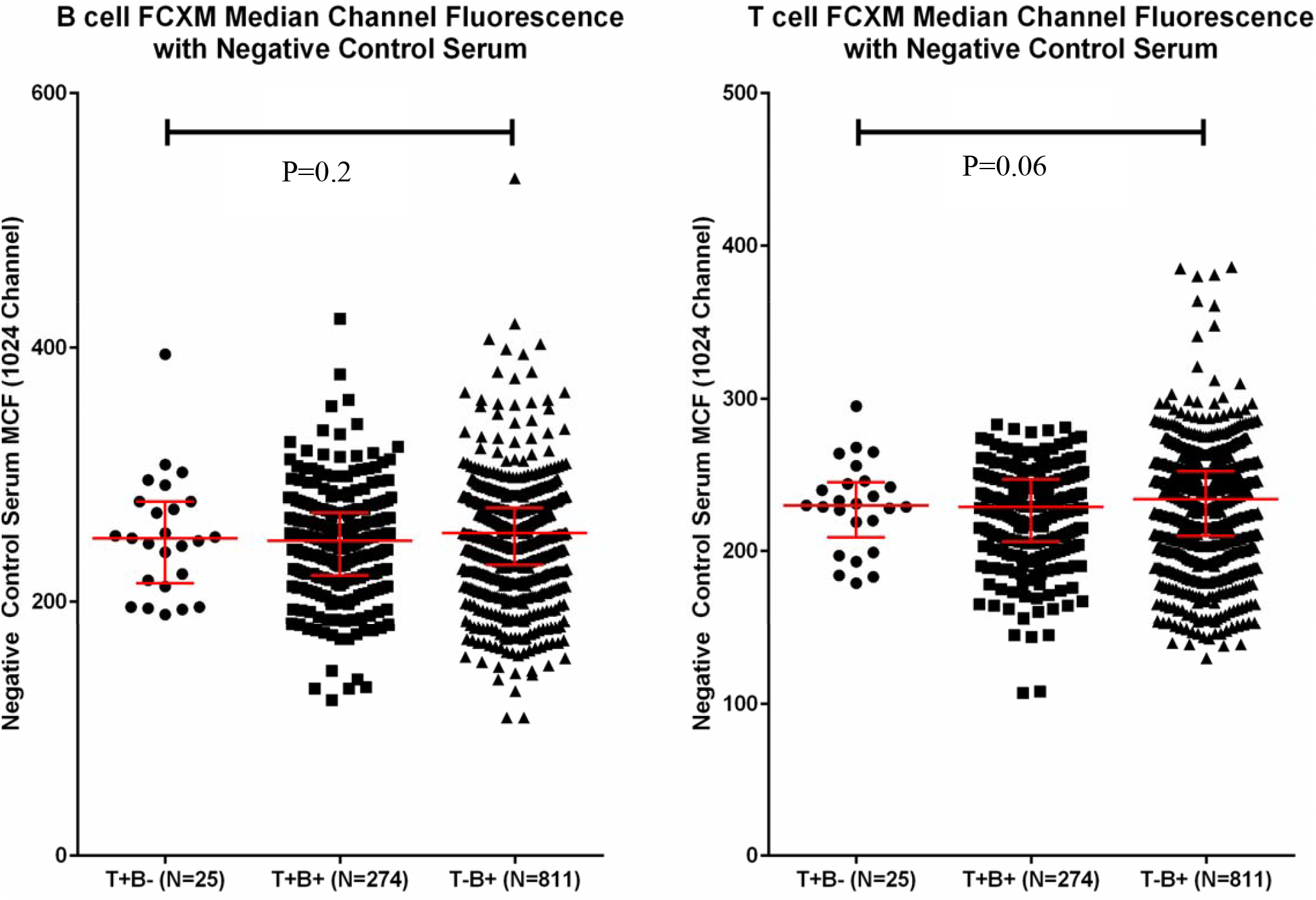
Distribution of T cell MCF and B cell MCF with cells treated with negative control serum. (A) Individual and median and 25^th^ percentile and 75^th^ percentile of MCF of B cells treated with the negative control serum are shown for the 25 T+B- FCXM tests, 274 T+B+ FCXM tests and the 811 T-B+ FCXM tests. (B) Individual and median and 25^th^ percentile and 75^th^ percentile of MCF of T cells treated with the negative control serum are shown for the 25 T+B- FCXM tests, 274 T+B+ FCXM tests and the 811 T-B+ FCXM tests. P values calculated using the Kruskal-Wallis test and included in each panel.

Another mechanism for the T+B- FCXM result is that the MCF of T cells treated with NCS was uniquely lower in T+B- FCXM compared to T+B+ or T-B+ FCXM. To investigate this possibility, we compared the MCF of T cells treated with NCS among FCXM scored as T+B-, T+B+ or T-B+. The median (IQR) of T cell MCF was 230 (36) in the 25 T+B- FCXM tests, 229 (41) in the 274 T+B+ FCXM, and 234 (43) in the 811 T-B+ FCXM (P=0.06) (Figure 3B). Thus, a uniquely low T cell MCF was an unlikely contributor to the T+B- FCXM.

A 50 CS is used as the threshold to classify a B cell FCXM as positive and a 40 CS to classify a T cell FCXM as positive in our laboratory. This differential cutpoint is a potential contributor to the T+B- FCXM result. Our data analysis showed that the 50 CS applied to score a B cell FCXM as positive may have contributed to 12 of 25 T+B- FCXM results (Table 1).

Figure 4 shows the flow cytometry acquisition setup and representative T+B- FCXM tests to illustrate potential mechanisms. The lymphocyte subpopulation was gated using FSC versus side SSC plot (Figure 4A). The T cells were gated using CD3 PerCP and the B cells using CD19 PE and the MCF of goat-anti-human IgG FITC fluorescence on T cells and on B cells were measured using histograms (Figure 4B). Figure 4C is a representative T+B- FCXM in which the 50 CS threshold used to score a B cell FCXM as positive contributed to the T+B-. Figure 4 D is a representative T+B- FCXM in which the 50 CS threshold used to score a B cell FCXM as positive did not contribute to the T+B- FCXM.

**Figure 4.**
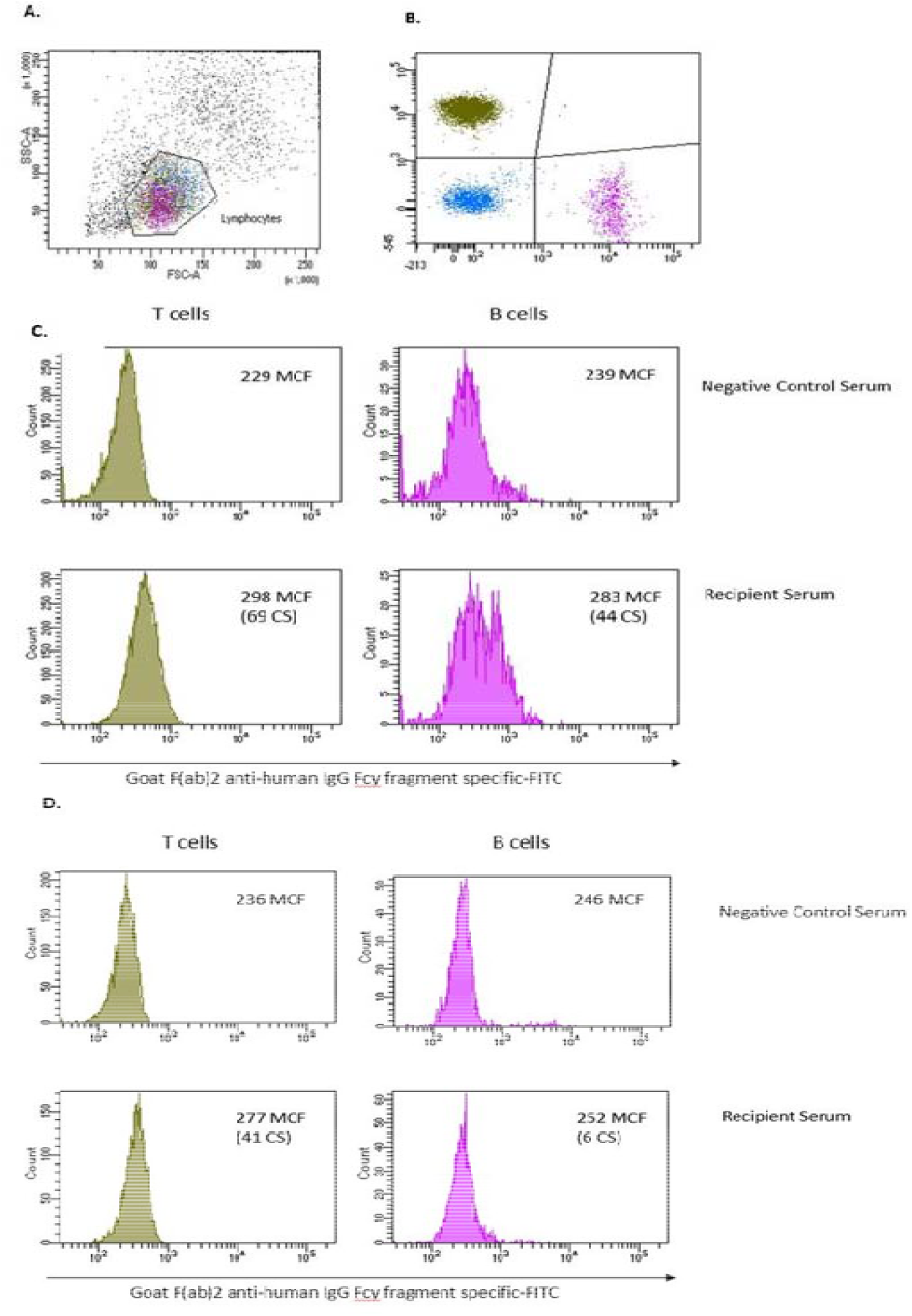
Flow cytometry acquisition setup and representative T+B– FCXM test results. (A) The lymphocyte subpopulation within the peripheral blood mononuclear cells were gated using forward scatter (FSC) versus side scatter (SSC) plot. (B) T cells (CD3 PerCP) and B cells (CD19 PE) were gated on the lymphocyte population. (C-D) Median channel fluorescence (MCF) of goat-anti-human IgG FITC fluorescence on T cells and on B cells were measured using histograms. Channel shift was calculated by subtracting the MCF with negative control serum from the MCF with recipient serum. (C) A representative T+B- FCXM test in which the 50 CS threshold used to score a B cell FCXM contributing to a T+B- FCXM result. Note a 50 CS to score the T cell as a positive test would still result in a T+B- FCXM. (D) A representative T+B- FCXM test in which the higher B cell CS threshold did not contribute to the T+B- test result. In Figure 4C-D, the MCF values are per 1024 channel scale values and the histograms are displayed in 5-decade log^10^ scale (FACS Diva software, versions 7.1.3 and 8.0.1).

### Clinical Outcomes

Three patients (Cases #16, #23 and #24) with a T+B- FCXM received kidney transplants at our center. All 3 received antithymocyte globulin as induction therapy and 2 (Cases # 16 and #23) received additional treatment with rituximab and intravenous immunoglobulin as per our transplant center policy for patients with DSA by LSA and a positive FCXM. The third patient, case #24 without DSA by LSA assay but a T+B- FCXM, did not receive additional therapy with rituximab or intravenous immunoglobulin. All three received maintenance therapy consisting of tacrolimus, mycophenolate mofetil and prednisone. None had an episode of acute rejection within the first year of transplantation and have excellent kidney graft function with serum creatinine levels of 0.88 mg/dL (Case #16), 1.0 mg/dL (Case #23) and 1.36 mg/dL (Case #24) at their last follow-up visit.

DSA monitoring showed that the DSA directed at B50 in Case #16 decreased from pretransplant MFI of 9349 to MFI of 1911 at 1-year post-transplantation. In Case #23, pre-transplant DSA MFI of 12350 remained at the same level and the MFI was 12418 at 1-year post-transplantation. Also, pre-transplant DSA MFI of 3673 directed at HLA-Cw17 increased to MFI of 5768 at 1-year post-transplantation. This patient also developed de novo DSA directed at HLA-B42 during this time period and the MFI was 4050. There were no pre-transplant DSA above MFI >2000 in patient # 24 and this patient did not develop de novo DSA during the first 3-months of following transplantation he was monitored.

## DISCUSSION

A systematic analysis of 3073 clinical FCXM performed using a standardized protocol in an ASHI-accredited Histocompatibility Laboratory identified that 8.4% of the T cell positive FCXM are T+B–. We found that antibodies directed not only at HLA-Cw but also antibodies directed HLA-A and antibodies directed at HLA-B are associated with a T+B– FCXM.

A mechanism for the T+B- FCXM test result is the differential expression of HLA-class I antigens on T cells and B cells. HLA-Cw antigen expression, as detected using mAb DT9, was found to be higher on T cells than B cells^12^, and in this study, seven of 10 FCXM T+B- FCXM were observed with sera containing HLA-Cw DSA. Sera with anti-HLA-Cw IgG had also been shown by others to cause T+B- FCXM^14,15^. Our findings are consistent with the earlier findings that antibodies directed at HLA-Cw antigens may contribute to a T+B- FCXM. Importantly, we report that antibodies directed at HLA-A and antibodies directed at HLA-B are also associated with a T+B- FCXM.

Our correlational analyses identified a significant association between the summed MFI of class I DSA and the MCF of T cells treated with the RS but not between the summed MFI of DSA and the MCF of B cells treated with RS. The basis for the lack of a relationship is unclear but may reflect aberrant expression of HLA class I on the B cells associated with a T+B- FCXM and impaired binding anti-HLA antibodies. The positive association between the summed MFI of class I DSA and the MCF of T cells treated with RS in this study was modest. The lack of a strong linear relationship may be related to the LSA assay being a qualitative assay rather than quantitative^16^ and possible confounding due to the presence of non-HLA antibodies. Our findings provide an explanation for the observation of Higgins et al^17^ that a serum with an MFI of 591 was associated with a positive FCXM whereas a serum with an MFI of 4773 was associated with a negative FCXM.

We reported that the MCF of B cells treated with NCS is significantly higher than the MCF of T cells treated with NCS^18^. The higher B cell MCF with NCS would reduce the B cell CS observed with RS and this could be a mechanism for the T+B- FCXM result. The higher B cell MCF however is unlikely to be the major mechanism since the MCF of B cells did not differ among FCXM that were classified as T+B-, T-B+ or T+B+ tests.

It is a common practice to use a larger CS to classify a B cell FCXM as positive compared to classifying a T cell FCXM as positive^19-22^. Had we applied the 40 CS threshold to classify not only T cell FCXM but also B cell FCXM, 12 of 25 T+B- FCXM would be reclassified as a T+B+ FCXM. Among these 12 FCXM tests, 7 of 12 would still be classified T+ FCXM if we had applied the 50 CS used for the B cell FCXM. Thus, the higher CS used to classify a B cell FCXM test is a potential contributor to only 5 of 25 T+B- results. In this regard, the CS threshold that should be applied to classify a FCXM test as positive or negative is the Achilles’s heel of this assay. In the FCXM, serum from a male donor of AB blood group and Rh negative and negative for anti-HLA-class I and class II IgG antibodies are used to establish baseline MCF of T cells and B cells and this serum is used as the NCS. The target cells are from heathy volunteers or potential donors. A predefined ratio of the fluorescence of the T cells and B cells treated with RS to the fluorescence of the corresponding cells treated with NCS is one approach to classify a FCXM^6^. A fluorescence value that is 2 or 3 standard deviations (SD) above the mean value observed with the NCS is also used to classify a FCXM as a positive or a negative^19-23^. A value that is 2 SD higher than the mean would place the observed value outside the 95% probability and a value that is 3 SD higher would place the observed value outside the 99.7% probability. However, this approach assumes normal distribution of fluorescence values. Fluorescence intensity is acquired using a logarithmic scale and is best estimated using median rather than mean, and the distribution of fluorescence values of T cells and B cells treated with NCS is seldom normally distributed. Indeed, the MCF values in the 3073 FCXM analyzed here were not distributed normally. These considerations led to the use of IQR value to derive CS in this study. Finally, had we used 2SD or 3SD instead of IQR to develop the CS, none of the 25 T+B- results would change.

In our study, we used the same CS for classifying FCXM irrespective of whether the cells were from a living donor or a deceased donor and regardless of whether the cells were isolated from peripheral blood, spleen or lymph node. The use of same CS irrespective of the donor cell type may be of concern since Badders et al. found that HLA-class I expression is lower on B cells isolated from deceased donor blood compared to B cells isolated from spleen, lymph node or B cells isolated from living donor blood^24^. The source of cells used as target cells in our study was not a significant issue since 22 of 25 T+B- were performed using PBMC.

Autoantibodies directed at T cells could result in a T+B- result. In our analysis of 1475 recipient auto FCXM performed in our laboratory during 2014 to 2018 (the same vintage as the 25 T+B- results reported here), not even a single auto FCXM was T+B- (0%) and 5 were T+B+ (0.34%), 119 were T-B+ (8.17%), and 1333 of 1475 were T-B-(91.49%). Thus, a T cell autoantibody being responsible for the observed T+B- FCXM appears extremely unlikely.

There are limitations to our study. The donor cells used in our FCXM tests were not treated with pronase, and our results may not be generalizable to tests performed using pronase treated cells. Pronase treatment has been reported to “improve flow cytometric detection of HLA antibodies”^25^ and “improve flow cytometry crossmatch results”^26^. However, pronase may also unmask cryptic epitopes on T cells^27^, elicit new reactivity to donor B cells and autologous B cells^28^, reduce HLA density on target cells leading to incorrect FCXM results^29^ including false positive T cell FCXM tests^30^.

The donors in our study were not HLA typed at high resolution and therefore the detected antibody may not be donor HLA allele specific. The LSA assay deployed in this study may not have included donor specific allele. These two limitations however apply to the lack of association between the summed MFI of DSA and MCF but not to the T+B- FCXM result since the B and T cells were from the same donor.

In sum, data analysis involving 3073 clinical FCXM tests performed using a standardized protocol in an ASHI accredited Histocompatibility laboratory identified that almost 10% of FCXM tests that are T+ FCXM are T+B- FCXM. We demonstrate that IgG antibodies directed at HLA-A, B and/or C locus determined antigens are associated with a T+B- result. Differential expression of HLA on T cells and B cells and/or differential binding of antibodies to T cells and B cells and CS used to qualify a FCXM test as positive or negative are potential contributors to the surprising T+B- FCXM result.

## Data Availability

Not applicable

## ACKNOWLEDGEMENTS

We gratefully acknowledge the expert technical help of our highly dedicated histocompatibility specialists from Immunogenetics and transplantation center, The Rogosin Institute.

## ABBREVIATIONS

ACD: Acid Citrate Dextrose
B cell: B lymphocyte
CD: cluster of differentiation
CDC-XM: complement dependent cytotoxicity crossmatch
CS: channel shift
DSA: donor specific antibody
EDTA: Ethylenediaminetetraacetic acid
F(ab’)_2_: fragment antigen-binding
Fc: fragement crystallizable
FCXM: flow cytometry crossmatch
FITC: Fluorescein isothiocyanate
FSC: forward scatter
HLA: human leukocyte antigen
IgG: Immunoglobulin G
IQR: interquartile range
LSA: Luminex single antigen
mAb: monoclonal antbody
MCF: median channel fluorescence
MFI: mean fluorescence intensity
NCS: negative control serum
PBMC: peripheral blood mononuclear cells
PBS: phosphate buffered saline
PCS: positive control serum
PE: Phycoerythrin
PerCP: Peridinin-Chlorophyll-Protein
RS: recipient serum
SSC: side scatter
T cell: T lymphocyte

